# Nigral volumetric and microstructural measures in individuals with scans without evidence of dopaminergic deficit

**DOI:** 10.1101/2022.10.26.22281257

**Authors:** Jason Langley, Kristy S. Hwang, Xiaoping P. Hu, Daniel E. Huddleston

**Affiliations:** Center for Advanced Neuroimaging, University of California Riverside, Riverside, CA, USA; Department of Neurosciences, University of California San Diego, San Diego, CA, USA; Department of Bioengineering, University of California Riverside, Riverside, CA, USA; Department of Neurology, Emory University, Atlanta, GA, USA

**Keywords:** neuromelanin, substantia nigra, SWEDD, Parkinson’s disease

## Abstract

**Introduction:** Striatal dopamine transporter imaging using 123I-ioflupane SPECT (DaTScan, GE) identifies 5-20% of newly diagnosed Parkinson’s disease (PD) subjects enrolling in clinical studies to have scans without evidence of dopaminergic deficit (SWEDD). These individuals meet diagnostic criteria for PD, but do not clinically progress as expected, and they are not believed to have neurodegenerative parkinsonism. Inclusion of SWEDD participants in PD biomarker studies or therapeutic trials may therefore cause them to fail. DaTScan can identify SWEDD individuals, but it is expensive and not widely available; an alternative imaging approach is needed. Here, we evaluate the use of neuromelanin-sensitive, iron-sensitive, and diffusion contrasts in substantia nigra pars compacta (SNpc) to differentiate SWEDD from PD individuals.

**Methods:** Neuromelanin-sensitive, iron-sensitive, and diffusion imaging data for SWEDD, PD, and control subjects were downloaded from the Parkinson’s Progression Markers Initiative (PPMI) database. SNpc volume, SNpc iron (R_2_), and SNpc free water (FW) were measured for each participant.

**Results:** Significantly smaller SNpc volume was seen in PD as compared to SWEDD (*P*<10^−3^) and control (*P*<10^−3^) subjects. SNpc FW was elevated in the PD group relative to controls (*P*=0.017). No group difference was observed in SNpc R_2_.

**Conclusion:** In conclusion, nigral volume and FW in the SWEDD group were similar to that of controls, while a reduction in nigral volume and increased FW were observed in the PD group relative to SWEDD and control participants. These results suggest that these MRI measures should be explored as a cost-effective alternative to DaTScan for evaluation of the nigrostriatal system.

## 1. Introduction

Neuronal loss in structures within the nigrostriatal system is a core feature of Parkinson’s disease (PD) [1, 2]. Much of this loss occurs in substantia nigra pars compacta (SNpc) during the prodromal stages of PD, and an estimated 30-50% of melanized neurons are lost by the time of symptom onset [2, 3]. Accordingly, efforts to develop neuroprotective therapies for PD increasingly focus on early-stage patients because the potential to prevent future neurodegeneration is greatest in this group. So far, no neuroprotective therapy has been established for PD at any stage [4], but early-stage PD patients have the most subtle features and can be especially challenging to diagnose. Thus, there is a pressing need for the development of imaging markers to assist selection of patients for recruitment into PD neuroprotection trials, particularly when these studies focus on early-stage PD patients. Imaging tools are also needed to monitor progression of neurodegeneration, and for development as surrogate outcome measures for these trials.

Striatal dopamine transporter imaging using ^123^I-ioflupane single photon positron emitted computed tomography (SPECT), also called DaTScan (GE Healthcare, Chicago, IL, USA), and ^18^F-fluorodopa positron emission tomography (PET) have found that between 5-20% of newly diagnosed PD subjects enrolling in clinical PD studies have scans without evidence of dopaminergic deficit (SWEDD) [5, 6]. Individuals with SWEDD meet clinical diagnostic criteria for PD but lack imaging evidence of nigrostriatal degeneration, the hallmark pathology of PD. A comparison of DaTScan images in a control participant, SWEDD participant, and PD participant is shown in Figure 1. Longitudinal study of SWEDD patients has found that they do not clinically progress significantly [7], and they are insensitive to treatment with levodopa [8]. Further, the highest percentage of SWEDD participants is observed in studies enrolling just after diagnosis [7, 9, 10]. While SWEDDs may be a heterogeneous group with a range of etiologies underlying their presentation with apparent clinical PD [5], they are unlikely to have idiopathic PD [7, 11] Therefore, inclusion of participants with SWEDD in biomarker studies or therapeutic trials for PD may cause them to fail due to inclusion of individuals without PD in the PD group.

**Figure 1.**
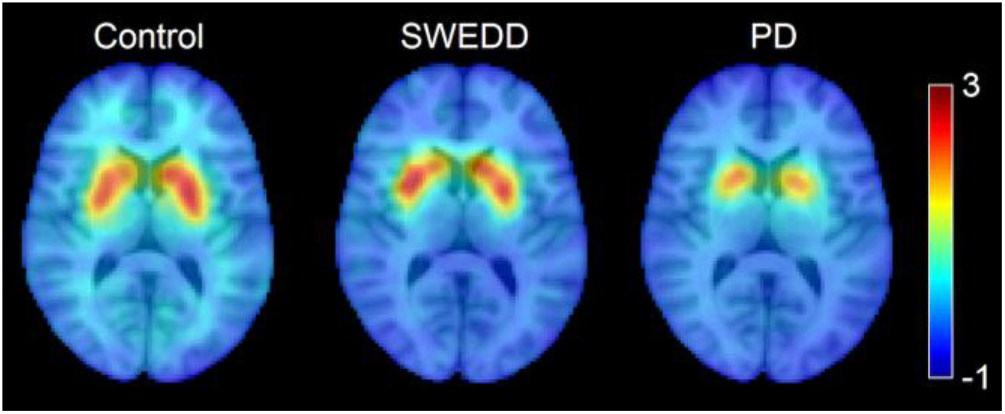
A comparison of striatal binding ratio derived from DaTScan images at the level of the striatum in a control participant (left), SWEDD participant (middle), and PD participant (right).

DaTScan has been used in numerous clinical trials and biomarker studies to confirm the presence of neurodegenerative parkinsonism. However, limited availability, high cost, substantial time requirement (a 3-6 hour delay is needed between radionuclide injection and scan), and safety concerns relating to the use of radioisotopes impedes the widespread adoption of DaTScan [12]. The scan-rescan reproducibility of DaTScan is also sub-optimal for quantitative analysis of DaTScan as an outcome measure [13]. An MRI-based imaging marker to corroborate the presence of neurodegenerative parkinsonism in individuals meeting diagnostic criteria for PD could improve biomarker study and trial designs in terms of cost-effectiveness, safety, and availability, and it may improve the odds of success of PD clinical trials.

Neuromelanin-sensitive MRI can evaluate dopaminergic neurodegeneration in SNpc *in vivo* without ionizing radiation. Neuromelanin-sensitive MRI images with explicit or incidental magnetization transfer (MT) effects allow quantification of neuromelanin-associated contrast in SNpc [14, 15] and MT contrast has been shown to colocalize with melanized neurons [16, 17]. Application of MT effects to examine nigral depigmentation has revealed PD-related reductions in nigral volume [18, 19] or SNpc area in a single slice [20]. Neuromelanin-sensitive MRI methods have also been shown to have high scan-rescan reproducibility [21-23].

Histological studies have observed iron deposition alongside nigral depigmentation in PD [24, 25]. MR relaxometry measures the relaxation rate of the transverse magnetization (R_2_ or R_2_*) and is sensitive to iron [26]. Increases in transverse relaxation rate in the substantia nigra regions of interest defined in T_2_- or T_2_*-weighted images [27-31] or in regions of interest defined by neuromelanin-sensitive contrast [32-36] have been found. The regions of interest placed by neuromelanin-sensitive contrast are likely placed in the SNpc since neuromelanin-sensitive contrast colocalizes with melanized neurons [16].

Diffusion MRI is sensitive to the diffusivity of water, and it allows researchers to probe tissue microstructure. Tissue microstructural changes in the setting of neurodegeneration likely include a local increase in extracellular fluid due to both cell loss and vasogenic edema caused by a neuroinflammatory response [37]. Bi-compartment models applied to diffusion MRI data to separate restricted diffusion in intracellular compartments from extracellular fluid, i.e., the free water (FW) compartment, have also been applied to examine PD neurodegeneration-related changes in SNpc. Increases in nigral FW have been observed in PD patients as compared to controls [38-41]. In this study, we use a multi-contrast approach to examine the nigrostriatal system in PD and SWEDD. Specifically, DaTScan is used to stratify the PD and SWEDD groups and to examine striatal dopamine transporter (DAT) uptake, magnetization transfer effects are used to examine nigral volume, relaxometry is used to examine iron deposition, and a bi-compartment model is used to examine nigral microstructure in PD patients, controls, and individuals with SWEDD.

## 2. Methods

### 2.1 PPMI Overview

Data used in the preparation of this article were obtained from the Parkinson’s Progression Markers Initiative (PPMI) database (www.ppmi-info.org/data). For up-to-date information on the study, visit www.ppmi-info.org. Full inclusion and exclusion criteria for enrollment in PPMI can be found at www.ppmi-info.org. Institutional IRB approved the study for each site and subjects gave written informed consent.

### 2.2 Participants

Explicit MT effects are generated by the application of fat saturation pulses prior to excitation [15] and incidental MT effects are generated by interleaved turbo spin echo (TSE) acquisitions [42]. These effects can be used to generate neuromelanin-sensitive images and evaluate the loss of neuromelanin-sensitive contrast in SNpc. The PPMI database was queried for individuals with T_1_-weighted MP-RAGE acquisitions, dual echo TSE acquisitions with a fat saturation pulse, and cardiac-gated diffusion tensor imaging (DTI) acquisitions.

Criteria for inclusion of subjects from the PPMI database used in this analysis were as follows: 1) participants must be scanned with cardiac-gated diffusion tensor imaging (DTI) and dual echo turbo-spin echo (TSE) with a fat saturation pulse and 2) participants must have DTI and TSE scans with scan parameters matching those in the PPMI imaging protocol. The 24-month time point was chosen for evaluation in the PD group since that time point contains the largest number of subjects with TSE acquisitions with a fat saturation pulse. A total of 163 subjects (33 controls (CO), 33 SWEDD, 97 PD patients) met these criteria. All PD and SWEDD subjects underwent DaTScan to confirm diagnosis. Imaging data were downloaded in December 2019.

### 2.3 MRI Acquisition

All MRI data used in this analysis were acquired on Siemens scanners. T_1_-weighted structural images in the PPMI cohort were used for registration to common space. Dual echo TSE images were acquired with the following parameters: TE_1_/TE_2_/TR=11/101/3270 ms, FOV=240×213 mm^2^, voxel size=0.9×0.9×3 mm^3^, fat saturation pulse, 48 slices. The first echo of the TSE acquisition contains magnetization transfer effects from the fat saturation pulse and interleaved TSE acquisition. Cardiac-gated diffusion-MRI data in the PPMI cohort were acquired using a monopolar diffusion encoding gradient with 64 unique gradient directions and the following parameters: TE/TR=88/650-1100 ms, flip angle=90°, FOV=229×229 mm^2^, voxel size=1.98×1.98×2 mm^3^, *b*=1000 s/mm^2^, cardiac-triggered, with 72 slices.

### 2.4 Image Processing

Diffusion data was preprocessed with FSL [43]. Diffusion MR data were corrected for motion and eddy-current distortions using EDDY in FSL. Next, susceptibility distortions were reduced by nonlinearly fitting the *b*=0 image to second echo from the TSE acquisition. Parameters derived from the diffusion data were estimated using DTIFIT in FSL. A bi-compartment model, as implemented in DIPY [44], was used to calculate free water images. Next, the *b*=0 image was brain extracted and registered to the brain extracted T_1_-weighted image using a rigid body transform with a boundary-based registration cost function.

*R*_2_, an MRI measure sensitive to iron, was calculated from the dual echo TSE acquisition using a custom script in MATLAB by fitting a monoexponential model to the dual echo TSE images. For each voxel, R_2_ was calculated from the dual echo TSE acquisition using the following equation:

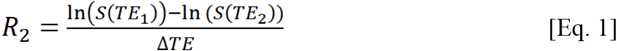

where *S*(*TE*_*i*_) denotes the signal at echo time *TE*_i_. The resulting *R*_2_ maps were aligned to the T_1_-weighted image using a rigid body transform derived via the magnitude image from the first echo.

### 2.5 Regions of Interest

An SNpc atlas, derived from magnetization-transfer images in a cohort of 76 healthy older participants (aged 66 years ± 6 years), was used as a region of interest (ROI) in the diffusion analysis [45]. The SNpc atlas was transformed from MNI space to subject space using linear and nonlinear transforms in FSL as previously described [46]. The SNpc atlas was thresholded at 60%, representing 60% of the population, and binarized. Mean FW and R_2_ was calculated in the SNpc for each subject.

SNpc was segmented using a thresholding method. A reference region was drawn in the cerebral peduncle in MNI common space and then transformed to individual NM-MRI images and used to threshold. Thresholding was restricted to the anatomic location of SNpc using the probabilistic standard space mask [45]. Voxels with intensity >μ_ref_+3σ_ref_ were considered part of SNpc.

### 2.6 Statistical Analysis

All statistical analyses were performed using IBM SPSS Statistics software version 24 (IBM Corporation, Somers, NY, USA) and results are reported as mean ± standard error. A *P* value of 0.05 was considered significant for all statistical tests performed in this work. A chi-squared test was used to evaluate gender differences in group. Normality of SNpc volumes and diffusion metrics was assessed using the Shapiro-Wilk test for each group and all data was found to be normal. The effect of group (SWEDD, PD, control) was tested with separate analysis of variance (ANOVA) for SNpc volume, mean SNpc FW, and mean SNpc R_2_. Since iron correlates with single-compartment diffusion indices, mean diffusivity and fractional anisotropy, were not used in this analysis [46]. ANOVA was used to test the effect of group (SWEDD, PD, control) on the ratio of caudate nucleus striatal binding ratio (SBR) to the putamen SBR. Caudate and putamen SBRs were obtained from summary variables in the PPMI database. For all ANOVAs, if the interaction was significant, post hoc comparisons between each pair of groups were performed using respective two-tailed t-tests. Correlations between SNpc volume, mean SNpc FW, and mean SNpc R_2_ and clinical measures (UPDRS and disease duration) were assessed in the PD and SWEDD groups. A correlation was considered significant if *P*<0.05 after multiple comparison correction. To test the feasibility of using MRI metrics to differentiate SWEDD from PD, receiver operator characteristic (ROC) curves were obtained for SNpc volume SNpc FW, and SNpc *R*_2_ values.

## 3. Results

### 3.1 Participants

Demographic data for each group is shown in Table 1. The ANOVA found no significant group effect in participant age (*P*=0.860; *F*=0.151), and education (*P*=0.322; *F*=1.41). No difference in gender was observed between PD and control groups (*P*=0.184), PD and SWEDD groups (*P=*0.747), and SWEDD and control groups (*P*=0.174). A significant group effect was seen in MoCA (*P*=0.042; *F*=3.218). The CO group had a higher MoCA score as compared to the SWEDD (*P=*0.019) and PD (*P*=0.034) groups. No difference was seen in MoCA between SWEDD and PD groups (*P*=0.439). A significant group effect was seen in UPDRS-III ON score (*P*<10^−4^; *F*=45.506) with the PD group having a higher UPDRS-III ON score than the CO (*P*<10^−4^) and SWEDD (*P*<10^−4^) groups. The SWEDD group had a higher UPDRS-III ON score as compared to the CO group (*P=*0.003).

**Table 1.**
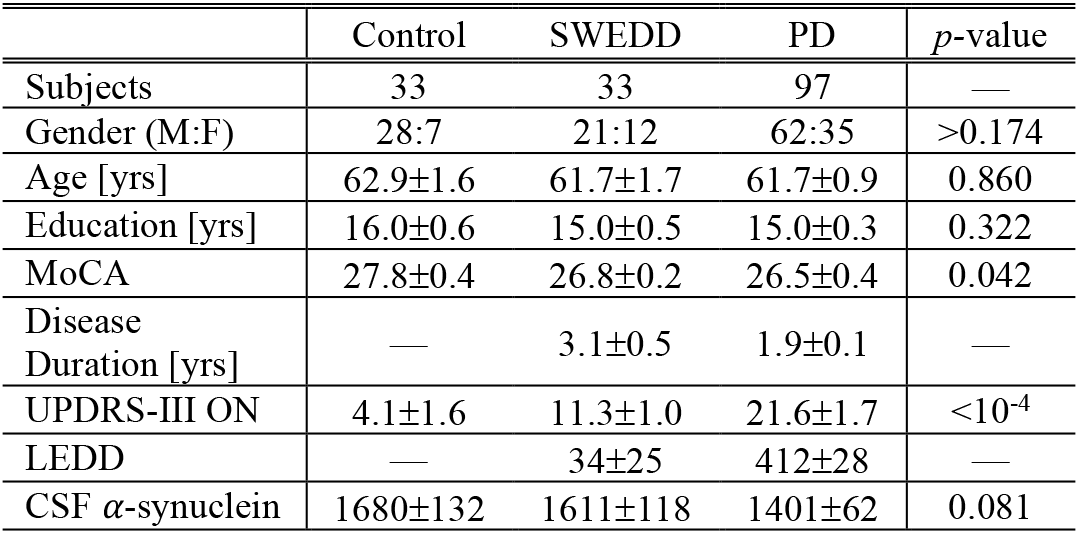
Demographic information for the groups used in this analysis. Data is presented as mean ± standard error. One-way ANOVAs were used for group comparisons of age, education, cognition, UPDRS-III ON medication score, and CSF *α*-synuclein from which *P* values are shown. A chi-squared test was used to assess group differences in gender.

|  | Control | SWEDD | PD | p-value |
| --- | --- | --- | --- | --- |
| Subjects | 33 | 33 | 97 | — |
| Gender (M:F) | 28:7 | 21:12 | 62:35 | >0.174 |
| Age [yrs] | 62.9±1.6 | 61.7±1.7 | 61.7±0.9 | 0.860 |
| Education [yrs] | 16.0±0.6 | 15.0±0.5 | 15.0±0.3 | 0.322 |
| MoCA | 27.8±0.4 | 26.8±0.2 | 26.5±0.4 | 0.042 |
| Disease Duration [yrs] | — | 3.1±0.5 | 1.9±0.1 | — |
| UPDRS-III ON | 4.1±1.6 | 11.3±1.0 | 21.6±1.7 | <10<sup>-4</sup> |
| LEDD | — | 34±25 | 412±28 | — |
| CSF α-synuclein | 1680±132 | 1611±118 | 1401±62 | 0.081 |

### 3.2 Effects of Group

A comparison of SNpc contrast in the first echo from the TSE acquisition for control, SWEDD, and PD subjects is shown in Figure 2. The effect of group (PD, SWEDD, control) was tested with ANOVAs in each MRI measure (SNpc volume, SNpc R_2_, free water). A significant main effect in group (*P*<10^−4^; *F=*33.475) was seen for SNpc volume. Pairwise-comparisons of means showed decreases in SNpc volume in the PD group (PD: 308 mm^3^ ± 14 mm^3^) relative to the control group (CO: 483 mm^3^ ± 24 mm^3^; *P*<10^−3^) and SWEDD (SWEDD: 493 mm^3^ ± 24 mm^3^; *P*<10^−3^) groups. No difference was seen in SNpc volume between SWEDD and control groups (*p*=0.235). No significant main effect was found in SNpc R_2_ (*P*=0.564; *F=*0.570). A significant main effect in group was seen in SNpc free water (*p*=0.027; *F=*3.710) with group means showing increases in SNpc free water in the PD group (PD: 0.198 ± 0.006) relative to control group (CO: 0.169 ± 0.010; *P*=0.017), and a non-significant trend toward increased SNpc FW in PD vs. SWEDD (SWEDD: 0.177 ± 0.010; *P*=0.074) was observed. No difference was seen in SNpc free water between the SWEDD and control (*P*=0.579) groups. Group comparisons are shown in Figure 3.

**Figure 2.**
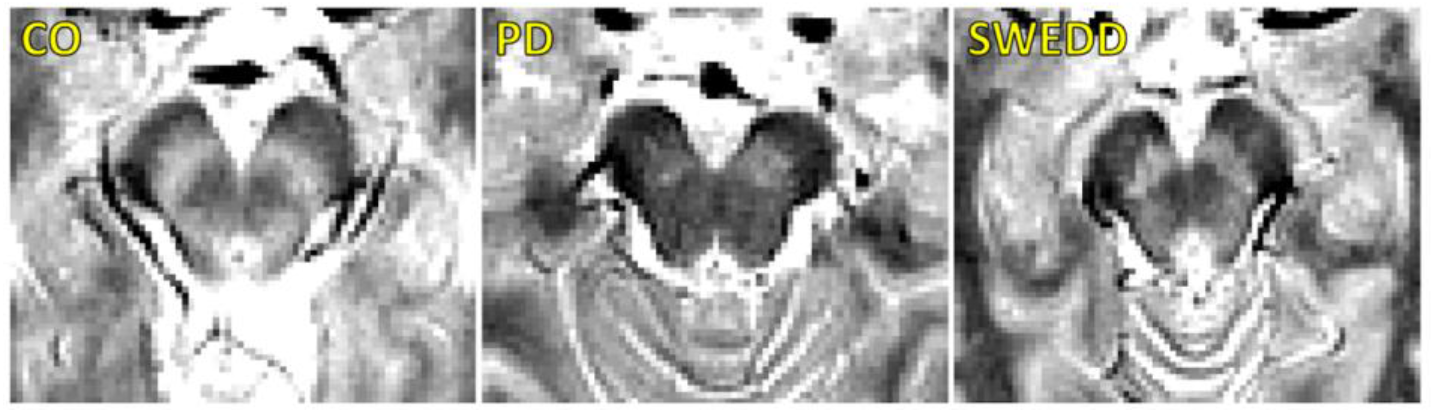
A comparison of neuromelanin-sensitive contrast from the first echo of the TSE acquisition in controls (left panel), PD (middle panel), and SWEDD (right panel) subjects. The PD subject exhibits a loss of contrast as compared to the control and SWEDD subjects

**Figure 3.**
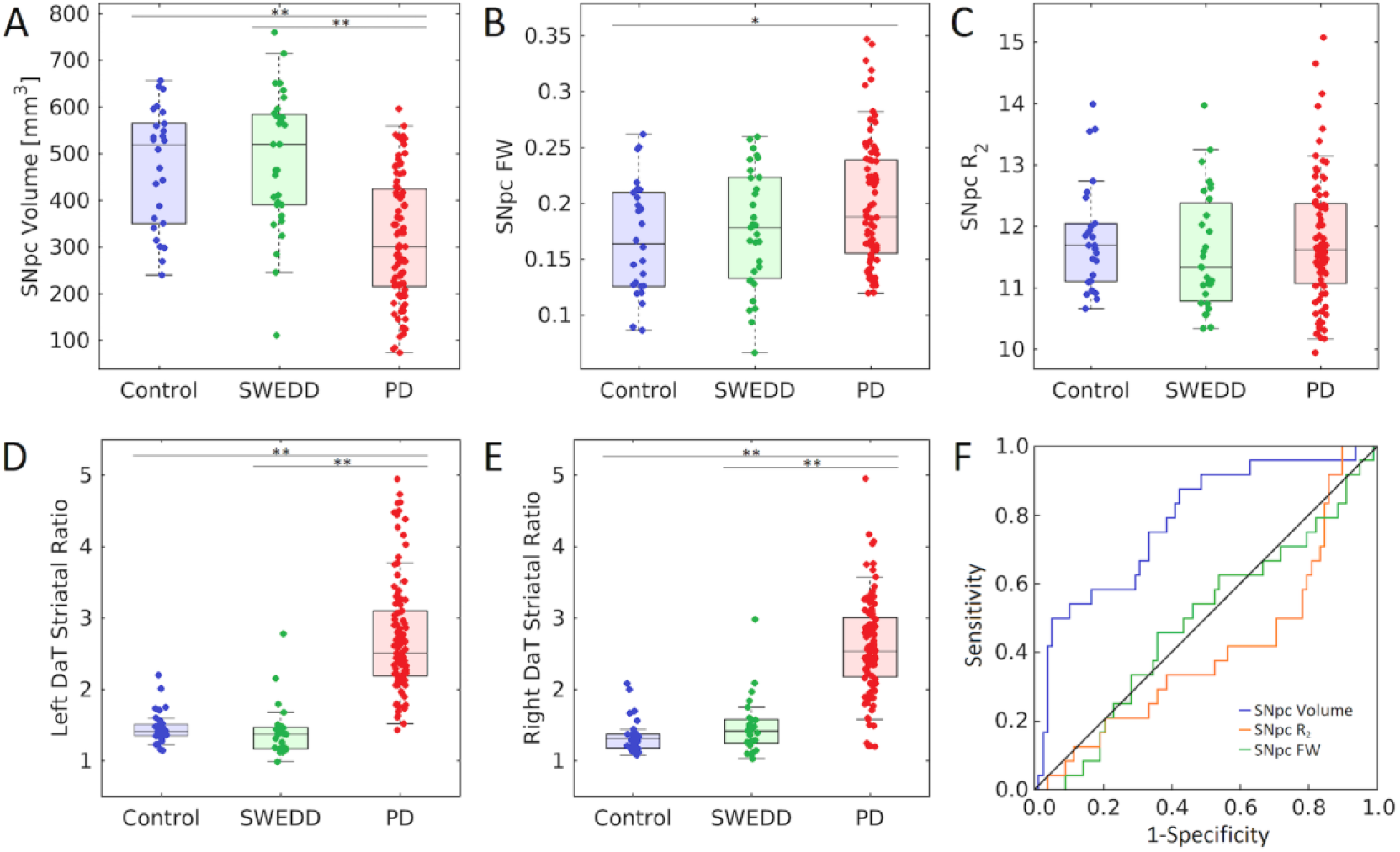
Group comparisons for SNpc volume (A), SNpc FW (B), SNpc R_2_ (C), left DaT striatal uptake ratio (D), and right DaT striatal uptake ratio (E) in control, SWEDD, and PD groups. AUC curves for SNpc volume, mean SNpc FW, and mean SNpc R_2_ are shown in (F). SNpc volume outperformed mean SNpc FW and mean SNpc R_2_ as a diagnostic marker to differentiate SWEDD from PD. All box plots, the top and bottom of the box denote the 25^th^ and 75^th^ percentiles, respectively, with the line denoting the median value. In (A-F), * denotes *P*<0.05 and ** denotes *P*<0.01.

Using the DaTScan data, the effect of group (PD, SWEDD, control) was tested with ANOVAs on striatal binding ratios (SBRs), i.e. the ratio of striatal uptake to occipital uptake. Specifically, the ratio of caudate nucleus SBR to putamen SBR was determined for each participant, and a significant main effect between groups was observed for the left (*F=*68.585, *P*<10^−3^) and right (*F*=41.538, *P*<10^−3^) ratios. Pairwise-comparisons of means showed increases in the ratio of caudate SBR to putamen SBR in the PD group (left: 2.7 ± 0.8; right: 2.7 ± 1.0) relative to control group (left: 1.5 ± 0.2; right: 1.3 ± 0.2) as well as relative to the SWEDD group (left: 1.4 ± 0.4; right: 1.5 ± 0.4) on both left and right sides (*Ps*<10^−3^). No difference was seen in the ratio of caudate SBR to putamen SBR between control and SWEDD groups (left: *P*=0.819; right: *P*=0.496).

### 3.2 Clinical Correlations

No significant correlations were observed between any MRI measure (SNpc volume, SNpc FW, or SNpc R_2_) and any clinical measure (UPDRS-III and disease duration) in the PD group (*Ps*>0.267) or in the SWEDD group (*Ps*>0.179).

### 3.2 ROC Analysis

SNpc volume differentiated SWEDD from PD better than SNpc FW or SNpc R_2_. The area under the ROC curve (AUC) for SNpc volume was 0.786 (standard error (SE)=0.048; 95% confidence interval (CI): 0.692–0.880; *P*<10^−4^). The AUC for mean SNpc FW was 0.489 (SE=0.060; 95% CI: 0.371–0.607; *P*=0.861) while the AUC for mean SNpc R_2_ was 0.405 (SE=0.060; 95% CI: 0.272–0.539; *P*=0.163). AUC curves are shown in **Figure 3**.

## 4. Discussion

Participants with SWEDD are individuals who have clinical features of parkinsonism and meet diagnostic criteria for PD, but they do not exhibit presynaptic dopaminergic deficits on DaTScan (see Figure 1). To date, the underlying pathophysiology in patients with clinical features of parkinsonism and SWEDD remains controversial. However, these individuals typically do not progress clinically or radiographically as expected in neurodegenerative parkinsonism [7], and they appear to represent a heterogeneous group that does not have idiopathic PD [11]. Currently, DaTScan is used as a screening tool to exclude SWEDDs in clinical PD studies. In research studies enrolling PD patients within 9 months of diagnosis approximately 5-20% of participants will be SWEDD, with the highest percentage occurring in studies recruiting newly diagnosed PD [7, 9, 10]. However, lack of availability of DaTScan, high cost, and exposure to ionizing radiation from the radioligand limit its use. Thus, there is a need for a widely available screening tool to exclude SWEDDs from enrollment in PD clinical studies, because these individuals are unlikely to have neurodegenerative parkinsonism [7]. This study examines the performance of nigral MRI metrics to differentiate SWEDD from PD. We found that nigral MRI metrics (SNpc volume, SNpc FW) in the SWEDD group resemble those derived in the control group and differ from PD.

The radiotracer used in DaTScan binds to dopamine transporter (DAT) in the presynaptic membrane of dopamine neuron axon terminals and is sensitive to the loss of membrane DAT protein, axonal neuronal loss, or nigrostriatal neuronal loss [47]. SWEDD patients do not show deficits in striatal DAT uptake, and our analysis of caudate nucleus to putamen SBR found results that were consistent with this. This is an expected result, since the PD and SWEDD groups are stratified based on their DaTScan results, but it is a useful confirmation of the basis of this study design. Neuromelanin-sensitive contrast is correlated with DAT binding [48, 49] and co-localizes with pigmented catecholamine neurons [16]. These results imply that SNpc should not undergo neurodegeneration in SWEDD. In agreement with this, no difference was observed in nigral FW and nigral volume between SWEDD and control groups.

In contrast to the SWEDD group, a reduction in SNpc volume was observed in the PD group relative to the control group. This result is in agreement with earlier studies that found nigral volume is reduced in PD patients [18, 19] or contrast is reduced in the lateral and ventral portion of SNpc [50]. The reduction in volume may be caused by depletion of melanized neurons in nigrosome-1, the subregion of SNpc that undergoes the greatest loss neuronal loss [1], as earlier studies have noted reductions in neuromelanin-sensitive contrast in regions consistent with nigrosome 1[15, 45]. Increases in nigral FW were also seen in the PD group relative to the control group, which is consistent with other studies reporting nigral FW increases in PD [39, 41].

When comparing SWEDD and PD, nigral volume outperformed nigral FW and nigral R_2_ as a diagnostic marker. These findings suggest that nigral volume, or other nigrostriatal imaging measures, could be useful for removal of SWEDDs in clinical trials. While the SNpc volume discrimination of PD vs. SWEDD approaches a useful level for clinical trial participant selection with an AUC of 0.786 (AUC > 0.8 is a typically discussed threshold), further improvements in accuracy are needed to make clinical decisions at the individual level, which would require AUC > 0.9. Diagnostic accuracy may be improved by using neuromelanin-sensitive acquisitions based on explicit magnetization transfer effects [22, 23] or incorporating other metrics, such as UPDRS score or MRI metrics from other structures, in machine learning classification algorithms. Metrics derived from neuromelanin-sensitive data show promise for differential diagnosis of parkinsonian syndromes [51-53] and further development of these metrics may allow for accurate discrimination of SWEDD from PD.

This study has several caveats. First, R_2_, a measure derived from a multiecho TSE sequence, was used to measure nigral iron deposition in PD and SWEDD. R_2_ is not as sensitive to iron deposition as R_2_* or susceptibility, which are derived from gradient-echo acquisitions, [26]. Further, R_2_* has been shown to be a robust marker for examining nigral iron deposition in PD [33] and may aid in differentiating SWEDD from PD. Second, incidental magnetization transfer effects from an interleaved multislice turbo spin echo acquisition were used to generate contrast sensitive to neuromelanin. Neuromelanin-sensitive MRI approaches employing explicit magnetization transfer effects exhibit high scan-rescan reproducibility in controls, outperforming methods based on incidental magnetization transfer effects [22, 23]. The use of explicit magnetization transfer effects may improve diagnostic accuracy for differentiating SWEDD from PD. Finally, no correlations were observed between MRI measures (nigral R_2_, nigral FW, nigral volume) and clinical features (disease duration, UPDRS-III score). The lack of correlation may also be explained in part by insensitivity of the imaging measures (incidental magnetization transfer effects and R_2_). However, since the PD subjects in PPMI are recruited at the time of initial diagnosis and therefore have very similar disease duration and clinical states (e.g., similar UPDRS-III scores), the lack of correlation may be due to insufficient variation in the clinical variables.

In conclusion, nigral volume and nigral FW in the SWEDD group were similar to that of the control group while a reduction in nigral volume and an increase in FW were observed in the PD group relative to the SWEDD and control groups. The changes in nigral volume and FW in the PD group are consistent with SNpc neurodegeneration effects in PD. With further development, nigral volume and FW may become markers used for differential diagnosis of SWEDD from PD. These imaging markers may be used to exclude individuals with SWEDD from PD neuroprotection trials.

## Data Availability

All data supporting these analyses are available through the Parkinson's Progression Markers Initiative (PPMI) database (www.ppmi-info.org/data).

## Acknowledgements

PPMI – a public-private partnership – is funded by the Michael J. Fox Foundation for Parkinson’s Research and funding partners, including [list the full names of all of the PPMI funding partners found at www.ppmi-info.org/about-ppmi/who-we-are/study-sponsors].

This work is supported by the NIH-NINDS 1K23NS105944-01A1 (Huddleston), NIH-NIA 1U19AG071754-01 (Huddleston, Hu, Langley), the Department of Veteran Affairs 1I01RX002967-01A2 (Huddleston), the Emory American Parkinson’s Disease Association Center for Advanced Research (Huddleston), the Emory Lewy Body Dementia Association Research Center of Excellence (Huddleston), and the Michael J Fox Foundation (MJF-10854, MJFF-010556; Huddleston, Hu, Langley).

